# Decreased thermal contrast sensitivity for paradoxical heat sensation in diabetic polyneuropathy

**DOI:** 10.64898/2026.11.25.26368869

**Authors:** Alexandra G. Mitchell, Jesper Fischer Ehmsen, Camilla E. Krænge, Arthur S. Courtin, Peter K. Brask-Thomsen, Sandra S. Gylfadottir, Francesca Fardo

**Affiliations:** Center of Functionally Integrative Neuroscience (CFIN), Department of Clinical Medicine, Aarhus University, Aarhus, Denmark; Center for Pain and Neuroplasticity (CNAP), Department of Health, Science and Technology, Aalborg University, Aalborg, Denmark; Institute of Neuroscience (IoNS), Université catholique de Louvain, Brussels, Belgium; Danish Pain Research Center (DPRC), Department of Clinical Medicine, Aarhus University, Aarhus, Denmark; Department of Neurology, Aarhus University Hospital, Aarhus, Denmark

**Keywords:** paradoxical heat sensation, diabetic polyneuropathy, thermal contrast, perceptual uncertainty, computational modelling

## Abstract

Paradoxical heat sensation (PHS), the perception of warmth during cooling, is typically induced by alternating warm and cold stimulation. Recent work has demonstrated that thermal contrast, defined as the difference between alternating temperatures, is a key determinant of this phenomenon in healthy individuals. However, it remains unclear whether peripheral sensory loss changes the thermal contrast required to elicit PHS and whether the experience itself remains the same after such sensory loss. We investigated PHS in individuals with probable diabetic polyneuropathy (DPN; N = 32) and healthy controls (N = 42) using a pre-registered Bayesian computational model. The model related the probability of a warm report to thermal contrast and used response times and confidence ratings as markers of perceptual uncertainty. Although prevalence was similar between groups, the thermal contrast required to elicit PHS was higher in patients with DPN, reaching the midpoint of the psychometric function at approximately 51% thermal contrast in healthy controls and 80% in patients with DPN. In both groups, warm reports were accompanied by greater uncertainty, reflected in longer response times and lower confidence. This pattern suggests that PHS does not typically reflect a clear sensation of warmth but rather an ambiguous thermal experience. These findings show that the sensation is accompanied by perceptual uncertainty in both groups, while patients with DPN require greater thermal contrast to elicit it.

## Introduction

Paradoxical heat sensation (PHS) refers to the perception of warmth or heat during cooling, typically induced by temporally alternating warm and cold stimulation of the skin. ^1–5^ Although unusual, PHS provides an opportunity to investigate how temporal patterns of thermal stimulation are transformed into perceptual experience. Recent work has demonstrated that thermal contrast, defined as the difference between the alternating warm and cold temperatures, is a key determinant of PHS perception in healthy individuals. As thermal contrast increases, so does the probability of experiencing PHS. ^6;7^

PHS has traditionally been considered uncommon in healthy individuals but frequently reported in patients with lesions of the peripheral or central somatosensory system, where it is associated with thermal sensory loss. ^8–14^ Recent findings, however, indicate that PHS is not confined to pathology. When thermal contrast is systematically manipulated, PHS can be observed in the majority of healthy individuals, ^6;7;15^ and its probability increases across the healthy adult lifespan. ^6^ These findings suggest that PHS may reflect a general property of thermosensory processing that emerges under specific temporal patterns of thermal stimulation. ^16^ On this view, its clinical association with thermal sensory loss could arise because this loss changes the conditions under which PHS is assessed, including the thermal contrast delivered during testing.

If PHS reflects a general thermosensory property, conditions that degrade thermal input provide a critical test. Diabetic polyneuropathy (DPN) affects peripheral sensory fibres, including the small-fibre pathways involved in warm and cold perception, resulting in impaired thermal sensitivity. ^17;18^ Increased PHS rates are reported in neuropathy, ^13;14^ and PHS has been identified as an early marker of thermal sensory loss over five years. ^14^ However, thermal contrast in these studies was determined by each individual’s detection sensitivity. Since reduced thermal sensitivity means larger temperature changes are required before detection occurs, observed differences in PHS prevalence in patients with DPN may reflect differences in the thermal stimuli between healthy and neuropathic individuals. This raises the question of whether increased PHS reported in patients with DPN simply reflects differences in the thermal contrast delivered during testing or altered PHS perception itself.

The present study aimed to characterise PHS perception in individuals with DPN and determine how it differs from healthy controls, using individually calibrated stimuli to manipulate thermal contrast. In addition, we used a pre-registered computational approach to investigate whether the relationship between thermal contrast and PHS differs in patients with DPN, as well as perceptual uncertainty surrounding the PHS experience. ^6^ Perceptual uncertainty refers to the noise or ambiguity in a sensory signal that makes a perceptual judgment less certain, ^19;20^ indexed behaviourally through response times and confidence. We recently used these measures to establish that PHS is an unusual perceptual experience for the majority of healthy adults. Our “unsure perceiver” profile showed that PHS responses were slower and less confident with increasing probability of experiencing PHS. ^6^ Here, we investigated whether individuals with DPN show the same perceptual profile as healthy controls of similar age.

## Methods

### Study design

The study employed a single-session, cross-sectional design. Participants completed three tasks aimed at assessing thermal sensitivity (detection and pain thresholding), ^21^ PHS ^6^ and spatial aspects of thermal perception. ^22^ In addition, all participants completed survey measures related to handedness, mental health, and pain sensitivity. The experiment took place at the Aarhus University Hospital, Aarhus, Denmark, and was approved by the Region Midtjylland Ethics Committee and adhered to the current version of the Declaration of Helsinki (2013). Participants provided informed consent prior to their participation in the study and were compensated for their participation.

This paper reports a pre-registered analysis of a subset of the dataset comparing PHS between patients with DPN and healthy controls. The analysis plan was pre-registered after completion of data collection but before any descriptive or inferential analyses of the patient data.

### Participants

The present analyses included 32 patients with probable DPN, recruited through the Danish Centre for Strategic Research in Type 2 Diabetes (DD2) cohort ^23–26^ and diagnosed according to the Toronto consensus criteria. ^17^ Probable DPN required two or more of the following: neuropathic symptoms, decreased distal sensation, or decreased or absent ankle reflexes, assessed by structured interview and bedside neurological examination. Additionally, 42 healthy controls of similar age were tested as part of an ageing cohort. ^6^

Of the 34 patients initially tested, two were excluded: one because of a previous thermal injury at the stimulation site, and one because of a technical error during temperature calibration. From the healthy cohort, nine participants were excluded from the whole cohort because they reported medications likely to influence thermal or pain perception. To match the patient sample, only healthy participants aged 47 years or older were included in the present analyses. General exclusion criteria included pregnancy or lactation, recent jet lag or sleep deprivation, non-diabetic skin disorders affecting the test site, a history of alcohol or drug abuse, cannabis use within the previous four weeks, and alcohol consumption within the previous 48 hours. For healthy controls, conditions or medications known to influence pain or thermal perception were additional exclusion criteria. The final sample comprised 74 participants.

Healthy controls had a mean age of 64.3 years (SD = 8.9; range 47–80; 17 men, 25 women), and patients with DPN had a mean age of 68.5 years (SD = 9.4; range 47–79; 17 men, 15 women).

### Stimuli and task

The apparatus, stimuli, and task were identical to those described in a previous study, ^6^ where full methodological details are provided. Stimulus intensities were individually calibrated from each participant’s warm detection and heat pain thresholds using the procedure described by Courtin et al. ^21^ All participants received task instructions in Danish.

Four peak temperatures used to manipulate thermal contrast were selected for each participant, corresponding to two innocuous (I-1, I-2) and two noxious (N-1, N-2) stimulation levels. Following Ehmsen et al., ^6^ these were derived from each participant’s warm detection and heat pain psychometric functions, estimated in a preceding thresholding task using the Psi adaptive Bayesian procedure. ^21;27^ I-1 corresponded to the warm detection threshold (50% detection probability) and I-2 to this threshold plus three times the inverse slope; N-1 corresponded to the heat pain threshold and N-2 to this threshold plus two times the inverse slope. The differences between I1 and I2 and between N1 and N2 were limited to a maximum of 4.5°C and 3°C, respectively. Peak temperatures were constrained to a safe range of 33–49.9°C. When the calculated I-1 or N-2 temperatures fell outside this range, they were adjusted to 33°C or 49.9°C, respectively. If four distinct ascending temperatures could not be generated within these limits, a predefined set of temperatures (34, 37, 44, and 45°C) was used. This occurred for two healthy controls and seven patients with DPN.

Thermal stimulation was delivered to the volar forearm using a T11 Thermal Contact Stimulator (QST.Lab, Strasbourg, France). Each trial began at a baseline temperature of 32°C. The thermode increased to one of the four individually calibrated peak temperatures at 2.5°C/s before returning to baseline at the same rate. A tone signalled the start of the critical cooling phase. Following a jittered delay (0.01-0.5 s), the thermode continued cooling below baseline at 1°C/s (**Figure 1A**). Participants pressed a button as soon as they detected the temperature change, which returned the thermode to baseline. They then indicated whether the sensation was cold or warm (PHS) and rated their confidence on a visual analogue scale ranging from guess (0) to certain (100) (**Figure 1B**).

**Figure 1.**
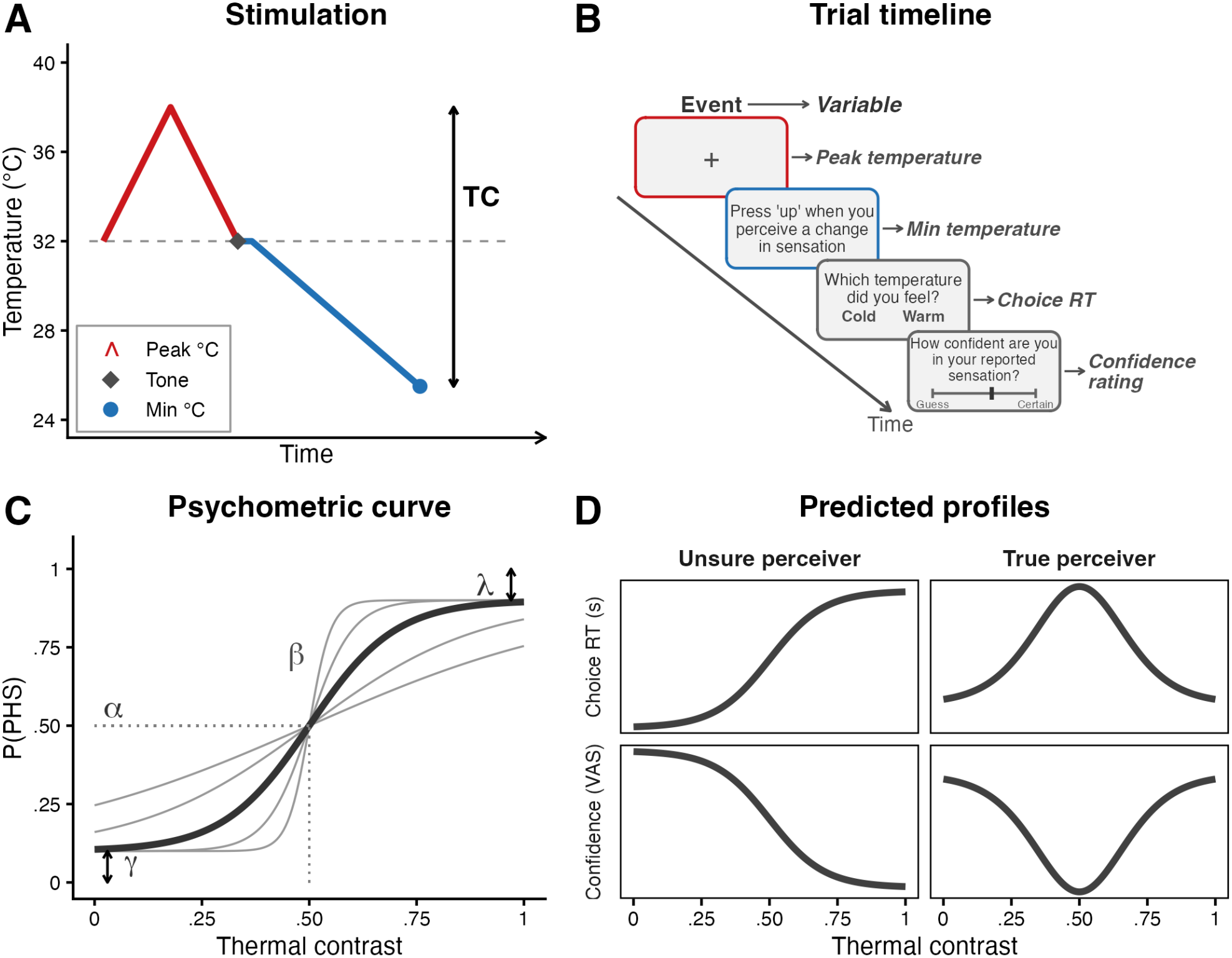
Task overview and theoretical framework. (A) Stimulation protocol for a single trial. The warm phase (red) ramps temperature above the 32 °C baseline and returns, followed by a cooling phase (blue) that decreases temperature below baseline. The thermal contrast (TC) is defined as the normalised difference between peak and minimum temperatures. (B) Trial timeline. Each trial began with a fixation cross during which the thermal stimulus was delivered (red border), followed by a detection phase in which participants pressed a button upon perceiving a change in sensation (blue border). Participants then reported the perceived temperature (cold or warm) and rated their confidence on a visual analogue scale (VAS) anchored from “Guess” to “Certain”. Arrows indicate the corresponding variables: peak temperature, minimum temperature, choice response time (RT), and confidence rating. (C) Schematic psychometric curve relating thermal contrast to the probability of reporting a paradoxical heat sensation (PHS). The curve is parameterised by threshold (*α*), slope (*β*, grey curves vary *β* only), guess rate (*γ*, lower asymptote), and lapse rate (*λ*, 1 *−* upper asymptote). (D) Predicted profiles for two hypothetical perceiver types. An unsure perceiver shows monotonically increasing choice RT and decreasing confidence with increasing P(PHS), reflecting growing uncertainty. A true perceiver shows non-monotonic patterns driven by decision entropy: both choice RT and confidence peak/dip near the psychometric threshold, where stimulus ambiguity is maximal.

Participants completed 64 trials, comprising 16 trials at each peak temperature. Two trials within each condition were catch trials in which the thermode warmed rather than cooled after the auditory cue, ensuring participants attended to the direction of the temperature change. Mean catch-trial accuracy was 90.6% (SD = 16.3) in healthy controls and 89.6% (SD = 15.1) in patients; no participants were excluded on this basis. To vary thermal contrast, half of the trials included a 1.5 s plateau at the peak temperature before cooling, whereas in the remaining trials cooling commenced immediately after the peak was reached.

### Thermal contrast

For each trial, we quantified thermal contrast using the Thermal Contrast Function (TCF) introduced by Mitchell et al. ^7^ The TCF expresses thermal contrast as the normalised difference between the warm and cold temperatures of a single trial, dividing this difference by the range between the maximum and minimum possible temperatures (50 °C and 0 °C). This provides a standardised value between 0 and 1, where higher values reflect a greater difference between the alternating warm and cold temperatures. Because the TCF summarises each trial’s warming and cooling in a single number, thermal contrast can be compared across trials, conditions, and groups on a common scale.

### Data analysis

All analyses and visualisations were conducted in R (version 4.4.2). ^28^ The hierarchical models were fitted in Stan ^29^ (CmdStan 2.35.0) via cmdstanr. ^30^ Analyses included calibration checks, computational modelling, classification analyses, and regression analyses. The pre-registered analysis plan is available at 10.17605/OSF.IO/QTVK2.

### Calibration checks and group comparisons

Peak temperature, minimum temperature, and thermal contrast were compared between groups within each peak temperature condition using Bayesian independent-samples t-tests with the default JZS prior. PHS prevalence was compared between groups using Bayesian contingency table tests (independent multinomial sampling with fixed group sizes, JZS prior). To examine PHS rates across peak temperature conditions, we fitted a Bayesian mixed-effects logistic regression with condition (4 levels) and group (HC vs. DPN) as fixed effects, and by-participant random intercepts and slopes for condition, with weakly informative priors (Normal(0, 1.5)) on all fixed effects. Three nested models were compared via bridge sampling to assess evidence for the condition *×* group interaction, the group main effect, and any group effect overall. Bayes Factors in favour of the null (BF_01_) were interpreted following conventional thresholds: BF_01_ > 3 as moderate and BF_01_ > 10 as strong evidence for equivalence; BF_01_ between 1/3 and 3 were considered inconclusive. These analyses were not included in our pre-registration but were added to investigate and formalise differences between patient and control groups.

### Computational modelling

We used a pre-registered multivariate Bayesian hierarchical model to jointly characterise PHS perception, response time, and confidence as a function of thermal contrast. The model followed the framework described by Ehmsen et al., ^6^ with full model specifications and detailed descriptions of the unsure perceiver and true perceiver formulations provided in that study. The model estimated how thermal contrast influenced the probability of experiencing PHS and associated behavioural markers of perceptual uncertainty.

PHS responses were modelled using a psychometric function with four participant level parameters (**Figure 1C**): threshold (*α*; the thermal contrast at the midpoint of the function, halfway between its lower and upper asymptotes), slope (*β*; the sensitivity of PHS probability to changes in thermal contrast), guess rate (*γ*; the lower asymptote of the function), and lapse rate (*λ*; one minus the upper asymptote of the function).

Response time was defined as the choice response time, that is, the time taken to indicate whether the sensation was cold or warm. Response times and confidence ratings were each modelled as an affine function of thermal contrast and a term reflecting perceptual uncertainty about the PHS percept. This uncertainty term distinguishes two hypothesised perceiver profiles (**Figure 1D**). The unsure perceiver links response time and confidence to the probability of PHS itself: choice RT increases and confidence decreases monotonically with the probability of PHS, as though a PHS percept is inherently uncertain. The true perceiver instead links them to the entropy of the PHS probability, which peaks when the probability of PHS is near 50%, where sensory evidence is maximally ambiguous, and decreases as evidence favours either percept: choice RT are slowest and confidence is lowest near the psychometric threshold, but both recover at high thermal contrasts, where sensory evidence is clear regardless of the percept reported. ^6^ The hierarchical model estimated individual differences in PHS sensitivity, response time, and confidence while accounting for correlations between these measures. Age and patient status (DPN vs. healthy control) were included as group-level predictors of the PHS threshold and of the response time and confidence intercepts.

Model estimation used Hamiltonian Monte Carlo sampling with four chains. Model convergence was assessed using R-hat statistics, effective sample size and absence of divergent transitions. ^31;32^ The four pre-registered models differed in whether response time and confidence were linked to the estimated probability of PHS (the unsure perceiver model) or to the entropy of that estimate (the true perceiver model), separately for each group within the same model (healthy controls vs patients with DPN). Bayesian inference was based on posterior means, 95% highest density intervals (HDIs), and the posterior probability that each parameter was greater than zero: i.e., *P*(*θ* > 0). Effects were considered credible when the 95% HDI excluded zero.

### Classification and further analyses

We assessed the diagnostic utility of PHS task parameters using a simulation-based ROC analysis with full uncertainty propagation from the hierarchical model posterior. For each of 1,000 simulations, we sampled group-level parameters from the posterior to generate synthetic patient and control datasets (N = 1,000 per group). A logistic regression classifier was trained on one simulated dataset and evaluated on an independent test dataset, with the parameters of the two datasets drawn from non-overlapping halves of the posterior draws and participant ages resampled from the observed sample. We compared a model based on PHS threshold alone with a model combining threshold, response time, and confidence ratings, with interactions between each parameter and age included in both models. Because the simulated datasets are generated from the fitted model, this analysis quantifies how well the estimated group differences separate patients from controls.

Additionally, to examine whether PHS is associated with thermal sensitivity ^21^ and spatial integration processes related to spatial summation and lateral inhibition, ^22^ we tested a further two pre-registered regression models. PHS threshold and slope were each regressed on cold and warm detection thresholds and psychometric slopes, as well as measures of spatial summation (i.e., the difference in perceived intensity between stimuli of different spatial extents) and lateral inhibition (i.e., the difference in perceived intensity between stimuli with identical overall area but different spatial separation). All predictors interacted with the group status (patients vs. controls) to test whether these relationships differed between groups.

## Results

### Thermal stimulation

Individually-calibrated peak temperatures, minimum temperatures reached during the cooling phase, as well as thermal contrast values showed no credible differences between groups in any of the four conditions. Bayesian t-tests provided inconclusive to moderate evidence for equivalence (peak temperature: BF_01_ = 2.81–3.98; minimum temperature: BF_01_ = 2.01–3.43; thermal contrast: BF_01_ = 1.52–3.75). Per-condition and per-group values are presented in **Table 1** and **Figure S1**. By design, I-1 corresponded to each participant’s warm detection threshold and N-1 to their heat pain threshold.

**Table 1.** Thermal stimulation and thermal contrast by condition and group.

|  | Peak (°C) |  | Minimum (°C) |  | Thermal contrast |  |
|---|---|---|---|---|---|---|
| Condition | HC | DPN | HC | DPN | HC | DPN |
| I-1 | 36.4 (2.8) | 37.1 (3.3) | 25.3 (4.4) | 23.6 (6.9) | .222 (.107) | .270 (.160) |
| I-2 | 39.9 (3.3) | 40.3 (3.5) | 24.7 (5.3) | 23.1 (7.3) | .304 (.136) | .343 (.171) |
| N-1 | 46.2 (2.2) | 46.0 (2.0) | 24.3 (5.5) | 23.3 (7.7) | .436 (.129) | .453 (.165) |
| N-2 | 47.7 (1.9) | 47.6 (1.9) | 23.9 (5.7) | 22.0 (8.1) | .476 (.127) | .511 (.174) |

*Note.* Values are group-level means (*SD*) across participants, computed from trials included in the computational model. HC = healthy controls; DPN = diabetic polyneuropathy.

### PHS prevalence

PHS prevalence, defined as reporting at least one PHS response across all trials, was observed in 38 out of 42 (90.5%) healthy controls and 29 out of 32 (90.6%) patients (**Figure 2A**), with moderate evidence for no group difference (BF_01_ = 5.84). At the stricter criteria (at least 5 to 30 PHS trials), evidence ranged from inconclusive to moderate (BF_01_ = 0.64–3.73). The mean number of PHS trials per participant was 15.3 (*SD* = 10.8) for healthy controls and 13.1 (*SD* = 9.9) for patients, and the mean per-participant percentage of PHS trials was 28.6% (*SD* = 20.4) for healthy controls and 24.1% (*SD* = 18.3) for patients. PHS rates were slightly higher on trials with a 1.5 s plateau (healthy controls: 29.2% vs 27.2%; patients with DPN: 25.9% vs 22.0%).

**Figure 2.**
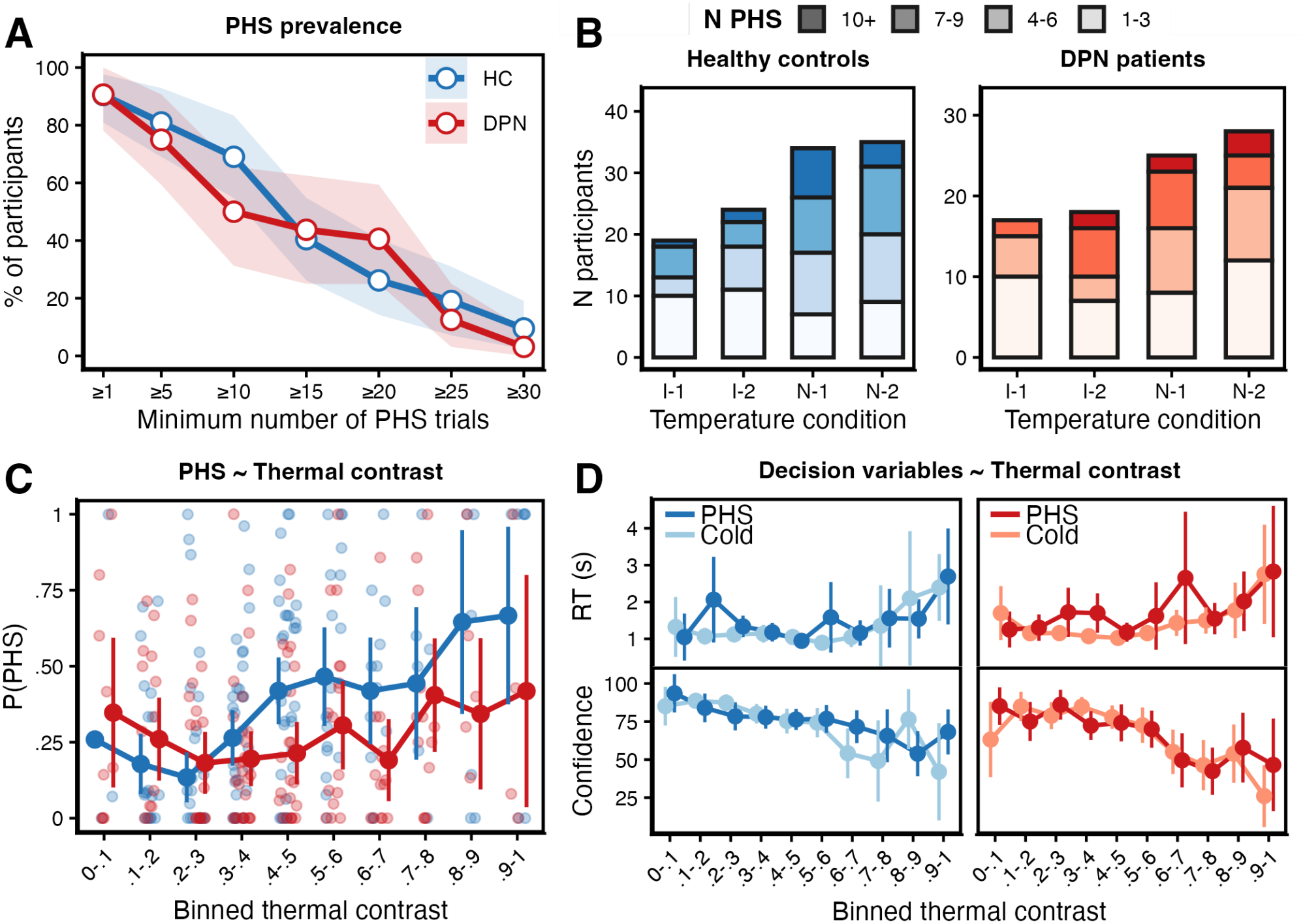
PHS prevalence, distribution, and raw decision variables. **(A)** Prevalence of paradoxical heat sensation (PHS) as a function of the minimum number of PHS trials per participant, shown separately for healthy controls (HC; blue) and patients with diabetic polyneuropathy (DPN; red). Shaded areas represent bootstrapped 95% confidence intervals (CIs). **(B)** Distribution of participants reporting PHS across peak temperature conditions (I-1 and I-2: innocuous; N-1 and N-2: noxious), stratified by group. Stacked bars indicate the number of PHS trials per participant (1–3, 4–6, 7–9, and *≥* 10). **(C)** Mean proportion of PHS trials per participant as a function of binned thermal contrast (0–1), shown separately for HC (blue) and patients with DPN (red). Individual dots represent participant-level means per bin; error bars indicate 95% CI. **(D)** Mean response time (RT; seconds; top) and confidence ratings (visual analogue scale [VAS]; bottom) as a function of binned thermal contrast, shown separately for PHS (dark) and non-PHS/cold trials (light) within each group (HC: blue shades, left; patients with DPN: red shades, right). Error bars indicate 95% CI.

To examine whether PHS varied as a function of peak temperature condition, we fitted a Bayesian mixed-effects logistic regression. PHS occurred more frequently during the noxious conditions (N-1 and N-2) than the innocuous conditions (I-1 and I-2). Bayesian model comparison provided strong evidence against a condition group interaction (BF_01_ = 11.77) and against any *×* group effect on PHS rates overall (BF_01_ = 12.48), indicating that PHS prevalence and its modulation by peak temperature condition were comparable between patients and healthy controls (**Figure 2B**).

### Thermal contrast, PHS, and perceptual uncertainty

To visualise how thermal contrast relates to PHS and to behavioural markers of perceptual uncertainty across groups, we divided thermal contrast into 10 equally spaced bins. For each participant we plotted the proportion of PHS trials per bin (**Figure 2C**), together with response times and confidence ratings across the same bins, separately for PHS and cold trials (**Figure 2D**). The computational model formalises these relationships as a psychometric function linking thermal contrast to the probability of PHS, with group-level parameters for threshold, slope, lapse and guess rates, and by linking response time and confidence to that probability estimate.

### The unsure perceiver model characterises PHS perception

We evaluated four models that specified different combinations of perceptual profiles across groups: unsure perceiver profiles in both patients with DPN and healthy controls, true perceiver profiles in both groups, and models in which the two groups differed in their assigned profile (unsure perceiver in patients and true perceiver in controls, or true perceiver in patients and unsure perceiver in controls). Only the model assuming an unsure perceiver profile in both groups met the pre-specified convergence criteria. The remaining models did not meet the pre-specified convergence criteria despite additional attempts to improve model estimation (see Supplementary Materials). Because only one model could be reliably estimated, the pre-registered model comparison could not be performed. Nevertheless, the descriptive response patterns were consistent with the unsure perceiver profile. Together, these findings indicate that the unsure perceiver model provided the most plausible account of PHS responses in the present sample, although this could not be formally confirmed through model comparison. All subsequent analyses were conducted using the unsure perceiver model for both groups.

### PHS threshold is increased in patients with DPN

The relationship between thermal contrast and PHS probability was characterised by four participant-specific parameters: threshold (the thermal contrast corresponding to the inflection point of the psychometric function), slope (governing the rate of increase in PHS probability with increasing thermal contrast), guess rate (the lower asymptote of the function, i.e. the probability of reporting PHS as thermal contrast approaches its lower limit), and lapse rate (one minus the upper asymptote, i.e. the probability of not reporting PHS as thermal contrast approaches its upper limit).

The group-level effect (patients with DPN vs. healthy controls) on the PHS threshold was credibly positive (log-scale estimate = 0.33, 95% HDI [0.12, 0.57], *P*(*θ* > 0) = .998; **Figure 3A**), corresponding to approximately 39% higher thresholds in patients at the average age and indicating that they required greater thermal contrast to reach the midpoint of the psychometric function. Averaged across participants, posterior mean thresholds were .51 (SD = .20) in healthy controls and .80 (SD = .29) in patients with DPN, corresponding to an approximately 30 percentage-point rightward shift. Observed thermal contrast ranged from .03 to .99 in healthy controls and from .04 to .98 in patients, so the estimated midpoints of both groups fell within the tested range.

**Figure 3.**
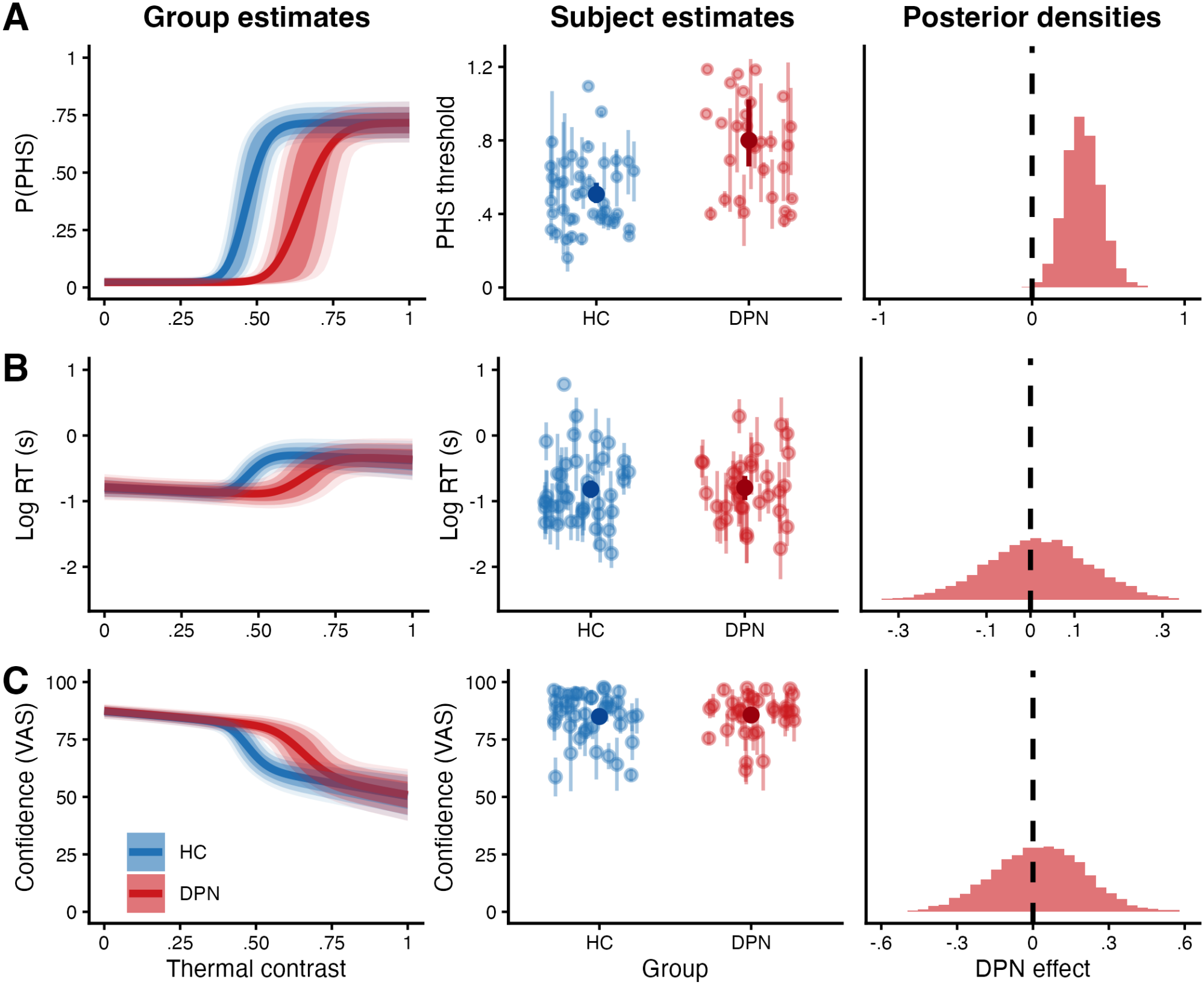
Computational modelling reveals increased PHS thresholds but preserved perceptual uncertainty in patients with DPN. Each row shows results for a different model parameter: **(A)** PHS probability, **(B)** log response time, and **(C)** confidence. Within each row, the left panel shows group-average model predictions as a function of thermal contrast for healthy controls (HC; blue) and patients with diabetic polyneuropathy (DPN; red), with shaded ribbons representing the 60%, 80%, and 90% posterior prediction intervals. The middle panel shows subject-level posterior mean parameter estimates by group (*±*68% credible intervals) with lighter colours, as well as group-level mean parameter estimates (*±*95% credible intervals) with darker colours: PHS threshold (**A**), log response time intercept (**B**), and confidence intercept (**C**). The PHS threshold represents the thermal contrast at the midpoint of each participant’s psychometric function (i.e., halfway between its floor and ceiling probability). The right panel shows the posterior distribution of the group effect (patients with DPN vs. healthy controls) on each parameter; dashed vertical lines indicate zero (no group difference). The threshold posterior (**A**) is credibly positive (95% HDI excludes zero), indicating a rightward shift of the psychometric function in patients with DPN. The response time (**B**) and confidence (**C**) posteriors overlap zero, indicating no credible group differences in these parameters.

Posterior predictive checks showed that the model reproduced the observed relationship between thermal contrast and PHS in both groups, at the group level and for individual participants (Figures S2–S5).

The psychometric slope (per unit thermal contrast on the 0–1 scale; HC: 148 *±* 116; DPN: 176 *±*181), guess rate (HC: .09 *±* .12; DPN: .15*±* .16), and lapse rate (HC: .27 *±* .07; DPN: .28 *±* .06; i.e. PHS probability reached a maximum of approximately .73 at the highest thermal contrasts) were numerically similar between groups, although these parameters were not formally compared. Patients with DPN therefore required greater thermal contrast to elicit PHS, with no indication of a change in the shape of the psychometric function.

### Higher PHS probability is associated with slower responses and lower confidence in both groups

PHS probability was positively associated with response time (mean = 0.93, 95% HDI [0.45, 1.40], P(*θ* > 0) > .999; Figure 3B), indicating slower responses when the estimated probability of PHS was higher. Conversely, PHS probability was negatively associated with confidence (mean = *−*1.41, 95% HDI [*−*2.02, *−*0.74], P(*θ* > 0) < .001; Figure 3C), indicating lower confidence when PHS probability was higher.

We found no credible evidence of a difference between patients with DPN and healthy controls in response time (mean = 0.02, 95% HDI [*−*0.22, 0.25], P(*θ* > 0) = .56) or confidence (mean = 0.02, 95% HDI [*−*0.33, 0.36], P(*θ* > 0) = .56), after accounting for thermal contrast and PHS probability. Thus, group differences in PHS were not accompanied by detectable differences in response time or confidence once PHS probability was taken into account.

### Classification of patients and controls using PHS parameters

PHS threshold discriminated patients from controls with modest accuracy (median AUC = .68, 95% CI [.55, .80], P(AUC*≤* .5) < .01). Adding response time and confidence ratings resulted in numerically similar classification performance (median AUC = .67, 95% CI [.53, .80], P(AUC *≤* .5) < .01). These findings indicate that the group difference is primarily reflected in the psychophysical threshold. Response time and confidence therefore provided limited additional information for diagnostic classification beyond the threshold estimate.

### PHS, thermal sensitivity and spatial integration processes

Of the 74 participants, 63 (36 healthy controls, 27 patients) had complete data across the three tasks of the overall study (i.e., thresholds, PHS; spatial integration); one healthy control lacked valid thermal thresholds, and ten participants (five controls, five patients) lacked spatial integration data. The regression model predicting PHS threshold from group, thermal detection thresholds and slopes, spatial integration measures, and their interactions with group explained a significant proportion of variance relative to an intercept-only model (F(13, 49) = 3.37, *p* = .001, R^2^ = .47, adjusted R^2^ = .33), indicating that the combined set of thermal detection thresholds and measures related to spatial integration explained substantial variance in PHS threshold. However, no individual predictor or group interaction reached statistical significance, suggesting that this explained variance was not attributable to a specific sensory measure or differential relationship between patients and controls. Variance inflation factors in a main-effects model were low (all ≤ 1.6). In contrast, the model predicting PHS slope did not explain significant variance relative to an intercept-only model (F(13, 49) = 0.99, *p* = .47, adjusted R^2^ *≈* 0), indicating that thermal sensitivity measures and spatial integration metrics did not predict how strongly PHS probability changed with increasing thermal contrast.

## Discussion

The present study shows that patients with diabetic polyneuropathy (DPN) require greater thermal contrast to elicit paradoxical heat sensation (PHS), without changing how PHS is experienced. Using a pre-registered Bayesian modelling approach, we identified three main findings. First, the unsure perceiver model was the only one of the four pre-registered models that converged, providing a plausible account of PHS in both patients and controls. This suggests that, for most participants, PHS is experienced as an uncertain and unusual sensory event rather than as a clear sensation of warmth. Second, patients with DPN required substantially greater contrast to experience PHS: the midpoint of the psychometric function was reached at approximately 51% thermal contrast in healthy controls and 80% in patients, reflecting a shift of the function. Third, response time and confidence showed similar patterns in both groups, suggesting that DPN affects the sensory signals giving rise to PHS rather than the uncertainty that accompanies it.

These findings extend those of Ehmsen et al., who used the same paradigm and computational framework across the healthy adult lifespan and found that the unsure perceiver model characterised PHS in younger and older adults. ^6^ The present study shows that this perceptual profile generalises to individuals with DPN. PHS was also common in both groups, with approximately 90% of participants reporting it on at least one trial and moderate evidence for no group difference. PHS is therefore not specific to neuropathy but reflects a general feature of thermosensory perception when warm and cold inputs are presented in close succession, accompanied by perceptual uncertainty whether or not the thermosensory system is affected by peripheral nerve pathology.

Despite this shared profile, the PHS threshold shifted in opposite directions in healthy ageing and in patients with DPN. Older adults required less contrast to experience PHS, reflected in a leftward shift of the psychometric function, ^6^ whereas patients with DPN required more, reflected in a rightward shift. In both cases the shape of the function was preserved, suggesting that ageing and DPN change the sensitivity of the thermosensory system while preserving its capacity to encode graded changes in contrast.

Ageing may lower the PHS threshold through a mild, predominantly cold-related decline in thermosensory function ^21^ or through changes in central processing, such as reduced cortical inhibition ^33^ or shifts in excitation–inhibition balance. ^34^ However, cold detection thresholds were not associated with PHS thresholds in our sample, and the present data cannot distinguish between existing accounts of PHS, such as disinhibition of warm-signalling pathways ^9^ or misattribution of degraded cold input to the warm channel. ^6^

In contrast, DPN may raise the threshold by weakening peripheral thermal signals. In the same participants, detection thresholds were elevated in patients with DPN, for warm (BF_10_ = 3.75) and, less conclusively, cold stimuli (BF_10_ = 1.76), whereas pain thresholds did not differ (BF_10_ *≤*0.34) (data from Courtin et al., ^21^ reanalysed for the present sample). Damage to small-fibre pathways attenuates thermal input, ^35^ so that greater contrast is required to generate equivalent sensory evidence at central processing stages. Together, these opposite shifts suggest that susceptibility to PHS may vary non-monotonically with thermosensory loss: partial decline, as in healthy ageing, may facilitate PHS, whereas more advanced loss of afferents, as in DPN, may weaken the signals from which PHS arises. This interpretation is consistent with clinical studies linking PHS to thermal hypoesthesia ^13^ and showing that it can precede measurable thermal sensory loss. ^14^

The comparable PHS prevalence between groups differs from clinical studies reporting higher PHS rates in individuals with neuropathy than in healthy controls ^14^ or DFNS reference values. ^10;13^ This discrepancy may reflect methodological differences. In the thermal sensory limen (TSL) procedure used in quantitative sensory testing, the thermal contrast experienced by participants is not experimentally controlled but emerges from individual detection thresholds. ^10;36^ Patients with impaired detection therefore experience larger temperature changes during testing and, because PHS probability increases with contrast, ^6;7^ are more likely to report PHS. In contrast, the present paradigm introduced systematic variability in contrast within each participant by combining four individually calibrated peak temperatures with a subsequent cooling phase, so that the relationship between contrast and PHS could be characterised across a range rather than at a single threshold-dependent value. Under these conditions, prevalence was comparable between groups, although patients required greater contrast to experience PHS, suggesting that neuropathy alters sensitivity to contrast rather than overall susceptibility to PHS. Differences in the number of trials between paradigms may also contribute to variation in reported PHS prevalence across studies.

The absence of group differences in response time and confidence suggests that DPN acts primarily on the sensory input underlying PHS rather than on its perceptual interpretation: peripheral nerve damage can change the relationship between stimulus and sensation without changing how uncertainty is experienced once a perceptual judgement is made.

Most previous research on DPN has focused on the structural and physiological consequences of small-fibre damage, including reduced intraepidermal nerve fibre density (e.g., ^37;38^), impaired peripheral nerve function (e.g., ^25;26;39^) and altered thermal thresholds (e.g., ^11^). Less is known about how peripheral nerve pathology shapes perceptual experience. Perception requires the nervous system to infer the most likely sensory state from incomplete and variable information, ^40;41^ and degraded peripheral input reduces the reliability of this evidence. Our findings extend this framework to thermosensation, showing that DPN alters the relationship between thermal contrast and PHS, consistent with less reliable sensory evidence.

Several limitations should be considered. First, the patient sample size was modest (N = 32), which may have limited sensitivity to detect small group differences in psychophysical parameters, response time and confidence. Second, only the unsure perceiver model met the pre-specified convergence criteria, so alternative models could not be formally compared using leave-one-out cross-validation; future work using larger samples and alternative model formulations may further characterise the mechanisms underlying PHS. Third, thermal stimulation was applied at a single site on the volar forearm, whereas DPN can affect body regions heterogeneously and typically affects the feet most severely. In addition, patients’ individual sensory profiles, such as the presence of neuropathic pain or predominantly cold or warm sensory loss, may have influenced sensitivity to thermal contrast; larger samples are needed to test whether PHS differs between sensory phenotypes. Finally, the cross-sectional design and the absence of stratification by neuropathy severity mean that the proposed non-monotonic relationship between thermosensory loss and PHS susceptibility could not be tested directly; longitudinal studies following individuals from incipient to established loss are needed.

## Conclusion

This study provides the first quantitative characterisation of the psychometric relationship between thermal contrast and PHS in individuals with DPN. Although overall PHS prevalence was comparable between patients with DPN and healthy controls, the thermal contrast required to elicit PHS was substantially higher in patients with DPN, who required greater thermal contrast to reach the midpoint of their psychometric function. This difference reflected a shift in threshold without a change in the shape of the psychometric function, suggesting that DPN reduces the strength of peripheral thermal signals while preserving the relationship between thermal contrast and PHS probability. Across both groups, the unsure perceiver model accounted for the key behavioural features associated with PHS perception, including slower responses and reduced confidence during PHS trials, supporting the characterisation of PHS as an inherently uncertain perceptual experience. These findings demonstrate the importance of distinguishing between PHS prevalence and sensitivity to thermal contrast and suggest that elevated PHS rates reported in neuropathy using standard clinical protocols may partly reflect the coupling between impaired thermal detection and PHS measurement. Modelling the stimulus–response relationship underlying PHS may therefore provide a complementary approach for characterising altered thermosensory processing in clinical populations.

## Data Availability

All data and analysis code produced in the present study are available online at https://github.com/Body-Pain-Perception-Lab/uncertainty-neuropathy-phs. Fitted model objects are available at https://osf.io/mkh4j/.

https://osf.io/mkh4j/

## Declarations

### Data and code availability

Data and analysis code are available at https://github.com/Body-Pain-Perception-Lab/uncertainty-neuropathy-phs. Model fits are available at https://osf.io/mkh4j/. The pre-registered analysis plan is available at https://doi.org/10.17605/OSF.IO/QTVK2.

### Funding

This work was supported by a European Research Council Starting Grant (ERC-2020-StG-948838) and a Lundbeck Foundation Experiment Grant (R436-2023-991) to FF.

### Conflicts of interest

The authors declare no conflicts of interest.

### Author contributions (CRediT)

Conceptualization: AGM, JFE, FF. Data curation: AGM, JFE, CEK. Formal analysis: AGM, JFE, FF. Investigation: AGM, CEK. Methodology: AGM, JFE, ASC, FF. Resources: PKBT, SSG. Software: AGM, JFE, FF. Supervision: FF. Funding acquisition: FF. Writing – original draft: AGM, JFE, FF. Writing – review & editing: all authors.

## Acknowledgement

We would like to thank Johanne Sejrskild Rejsenhus for her help with data collection and Dr. Camila Sardeto Deolindo for her contribution to the thermal thresholding procedure. We also thank all participants, including those recruited from the Danish Centre for Strategic Research in Type 2 Diabetes (DD2) cohort.

## Notes

### Competing Interest Statement

The authors have declared no competing interest.

### Clinical Protocols

https://doi.org/10.17605/OSF.IO/QTVK2

### Author Declarations

The Committees on Health Research Ethics for the Central Denmark Region (De Videnskabsetiske Komiteer for Region Midtjylland) gave ethical approval for this work (case no. 1-10-72-168-23).

