## Supplementary Materials for "Decreased thermal contrast sensitivity for paradoxical heat sensation in diabetic polyneuropathy"

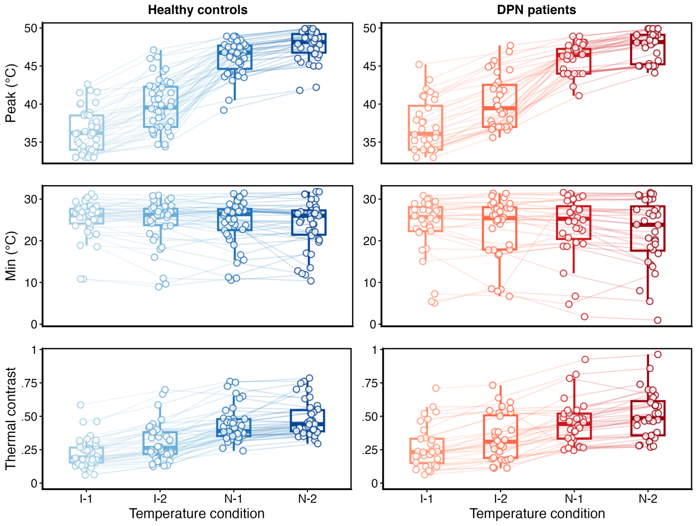


**Figure S1. Thermal stimulation parameters by temperature condition and group.** Individual peak temperatures (top), minimum temperatures (middle), and thermal contrast (bottom) across the four temperature conditions (I-1, I-2: innocuous; N-1, N-2: noxious peak temperatures) for healthy controls (HC, left, blue) and diabetic polyneuropathy patients (DPN, right, red). Boxplots show the median and interquartile range. Open circles represent individual participant means per condition; lines connect the same participant across conditions. Colour shading progresses from light to dark with increasing condition intensity.

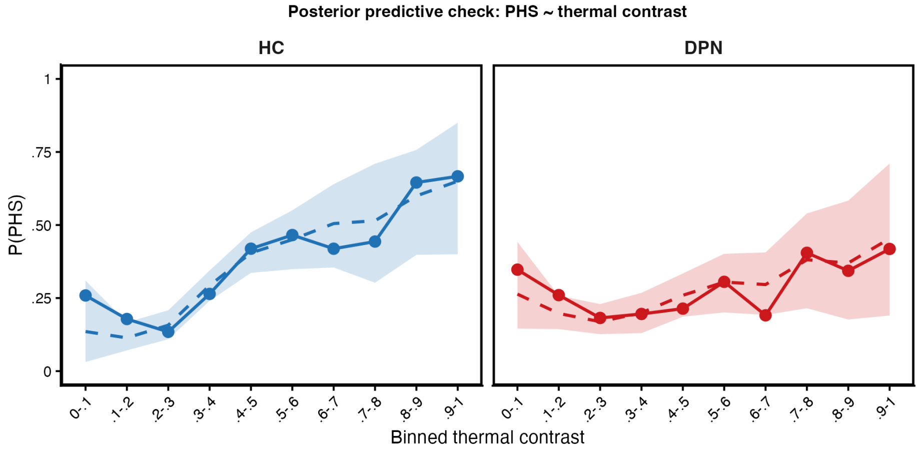


**Figure S2. Posterior predictive check for the relationship between thermal contrast and PHS (cf. Figure 2C).** Points and solid lines show the observed mean proportion of PHS trials per binned thermal contrast (participant means per bin, averaged within group) for healthy controls (HC, blue) and patients with diabetic polyneuropathy (DPN, red). Dashed lines and shaded areas show the median and 95% interval of the same summary computed from 1,000 datasets simulated from the posterior of the unsure perceiver model.

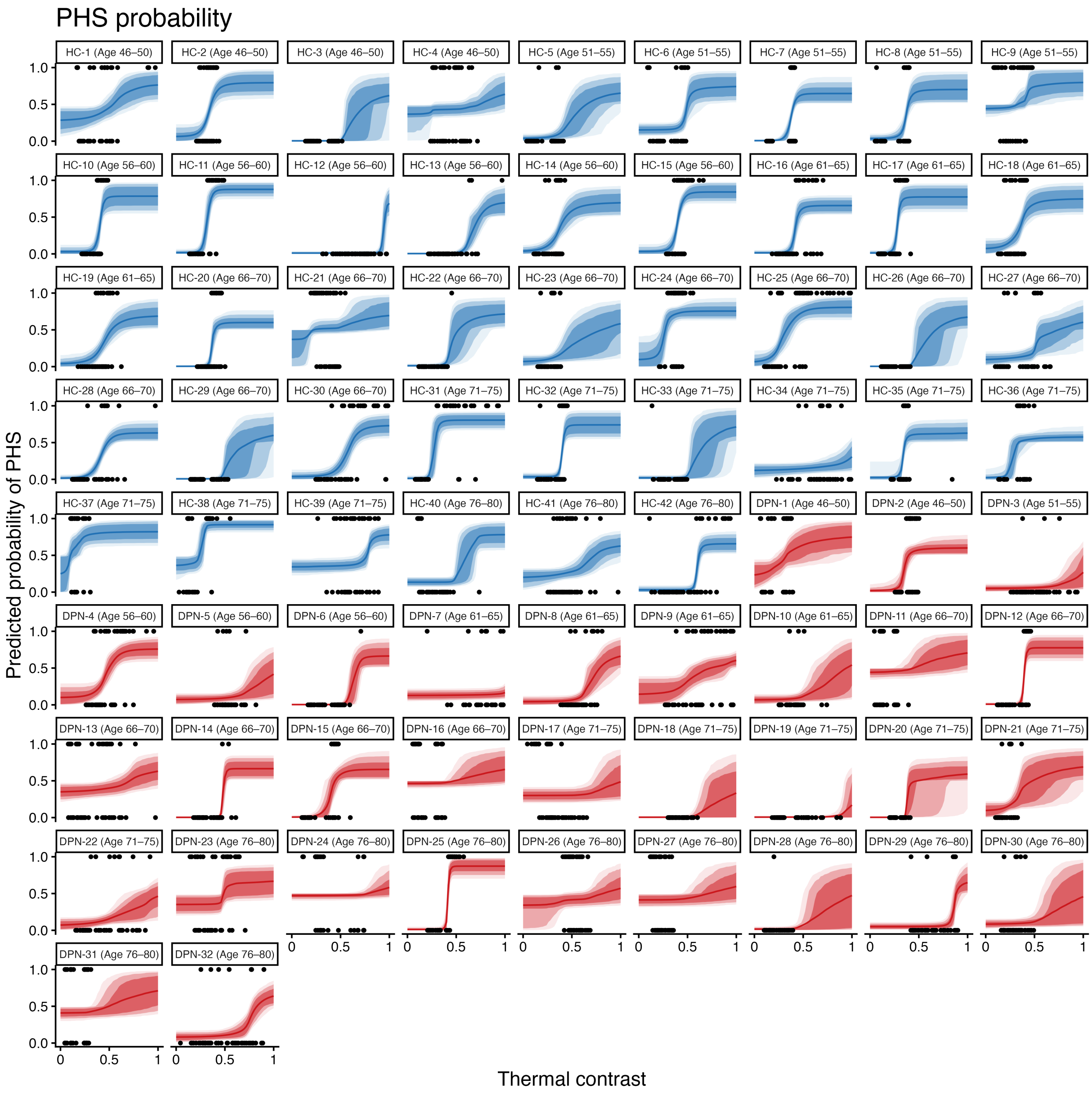


**Figure S3. Binary PHS responses and posterior estimates of predicted PHS probability as a function of thermal contrast for each participant, ordered by group (healthy controls, HC, blue; patients with diabetic polyneuropathy, DPN, red) and by age within group; labels give 5-year age bands.** Solid line shows the mean estimate; shaded bands show 90%, 80% and 60% credible intervals.

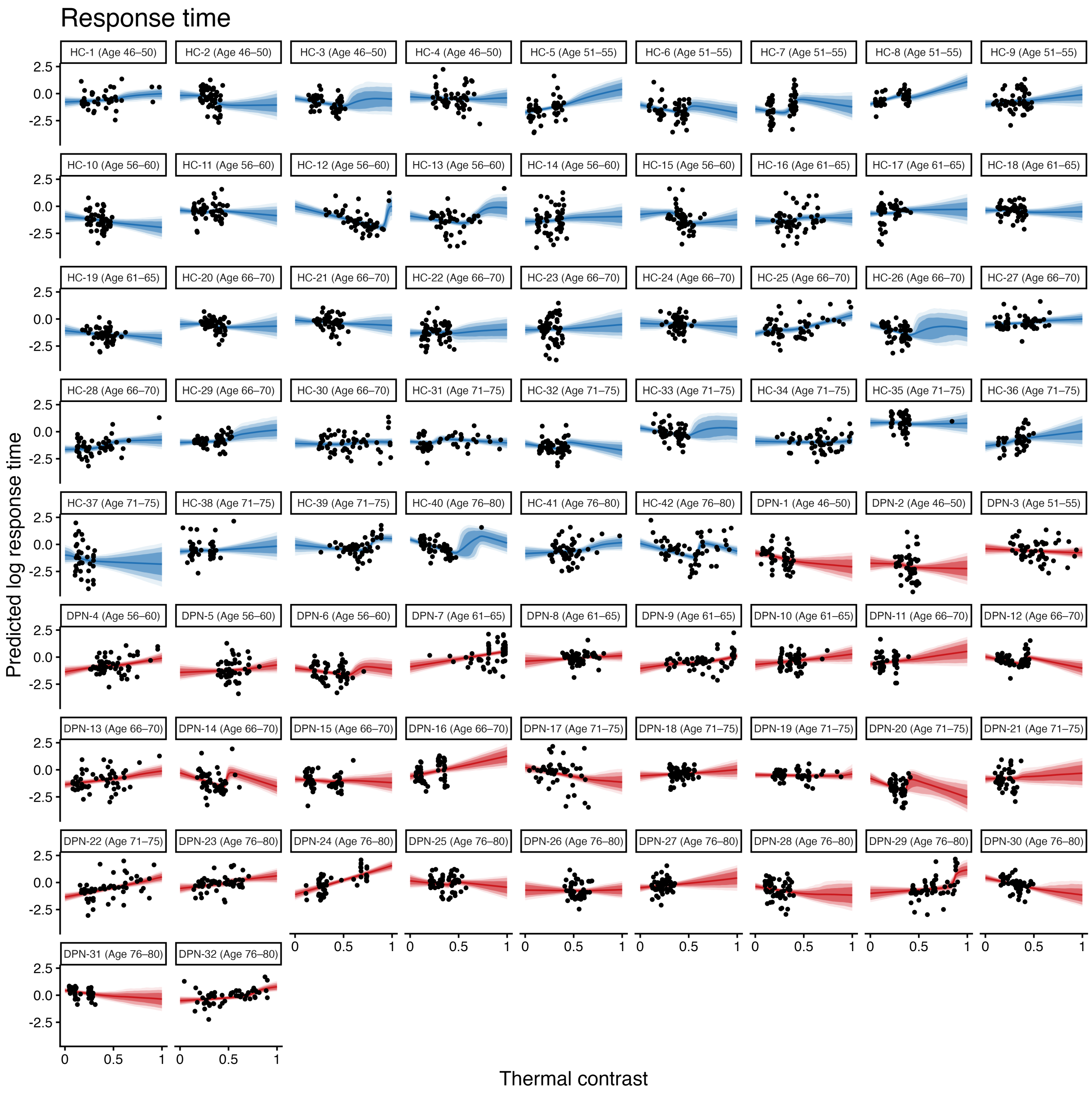


**Figure S4. Log response times (minus the estimated non-decision time) and posterior estimates of mean log response time as a function of thermal contrast for each participant, ordered by group and by age within group; labels give 5-year age bands.** Solid line shows the mean estimate; shaded bands show 90%, 80% and 60% credible intervals.

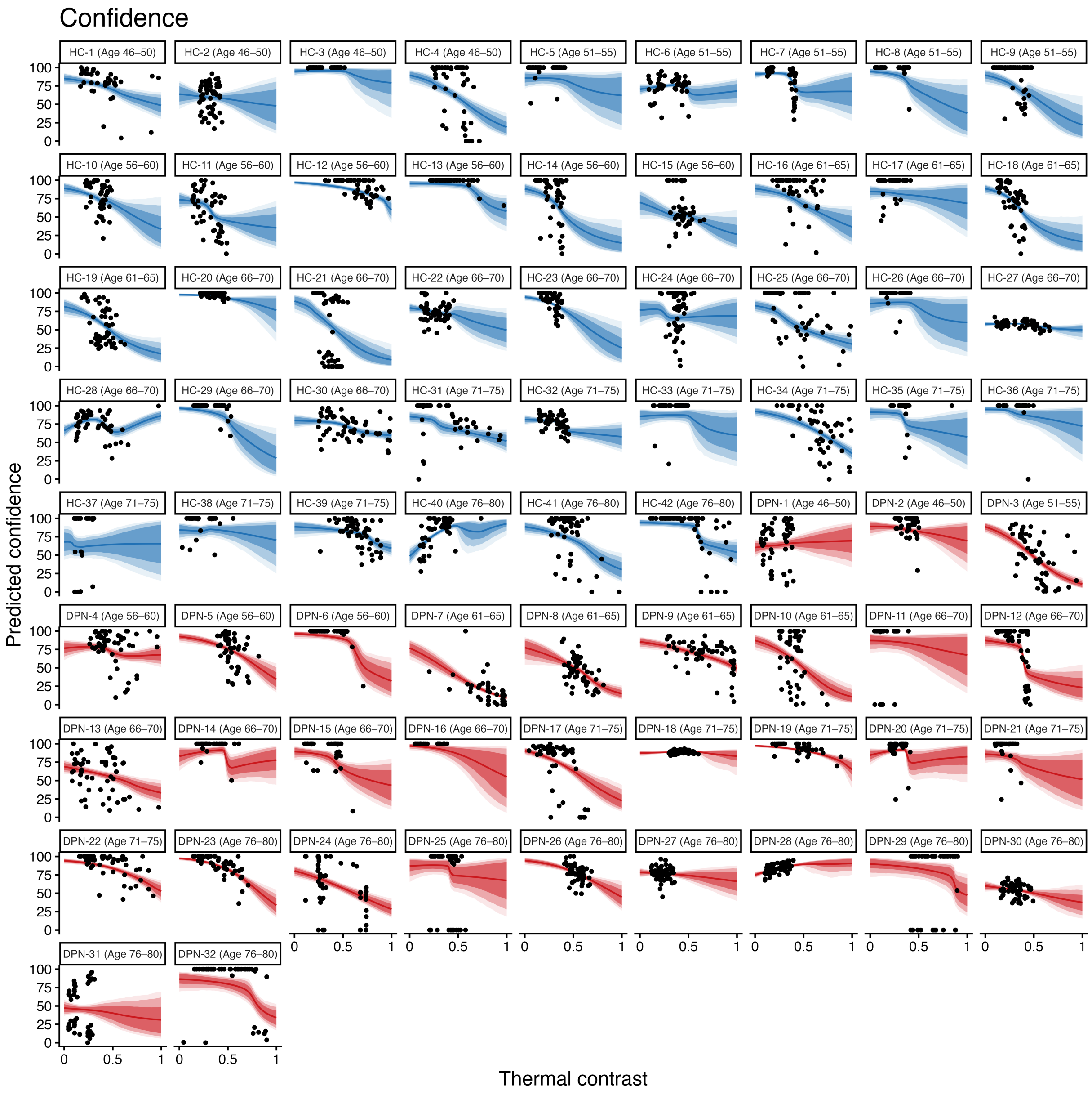


**Figure S5. Confidence ratings and posterior estimates of mean confidence as a function of thermal contrast for each participant, ordered by group and by age within group; labels give 5-year age bands.** Solid line shows the mean estimate; shaded bands show 90%, 80% and 60% credible intervals.

### Deviations from the pre-registration

The pre-registration is available at https://doi.org/10.17605/OSF.IO/QTVK2. Deviations are reported below.

- *Planned model comparison:* The pre-registered leave-one-out cross-validation could not be performed because only one of the four pre-registered models met the convergence criteria. Consequently, all subsequent analyses were based on the only estimable model (the unsure perceiver model).
- *Additional analyses:* Calibration and group comparisons of thermal stimulation parameters, and group comparisons of PHS prevalence, were not pre-registered. These analyses were conducted to provide descriptive context for the computational modelling results.
- *Priors:* Priors for the PHS psychometric parameters, age effects and group differences were as pre-registered. Priors for the response-time and confidence submodels were misreported in the pre-registration; the priors used are listed in Table S1.
- *Convergence criteria:* The pre-registration specified that no inference would be made on models with divergent transitions or with group-mean R-hat values above 1.01. The reported model had 3 divergent transitions out of 8,000 post-warm-up iterations, and the group-level threshold intercept had an R-hat of 1.012; all other group-level parameters met the criteria (bulk ESS $\geq$ 514). Following the pre-registered contingency plan, the maximum tree depth was increased from 12 to 15.

### Prior distributions

**Table S1.** Prior distributions of the reported (unsure perceiver) model.

| Parameter | Scale | Group mean | Between-participant SD |
|---|---|---|---|
| **PHS** | | | |
| Threshold | log | N(4, 0.5) | N+(1, 0.5) |
| Slope | log | N(−2, 1.5) | N+(0, 1) |
| Guess rate | logit | N(−5, 1) | N+(0, 1) |
| Lapse rate | logit | N(0, 1) | N+(0, 1) |
| **Response time** | | | |
| Intercept | log RT | N(−1.5, 0.5) | N+(1, 1) |
| Effect of P(PHS) | log RT | N(1, 0.5) | N+(1, 2) |
| Effect of thermal contrast | log RT | N(0, 0.005) | N+(0, 0.5) |
| Residual SD ($\sigma$) | log | N(−1, 2) | N+(1, 1) |
| Non-decision time | logit | N(0, 1) | N+(1, 1) |
| **Confidence** | | | |
| Intercept | logit | N(0.9, 0.5) | N+(0.5, 1.5) |
| Effect of P(PHS) | logit | N(−2, 1) | N+(0, 1) |
| Effect of thermal contrast | logit | N(0, 0.005) | N+(0, 0.5) |
| Precision ($\phi$) | log | N(2, 1) | N+(1, 1) |
| Cut-point 0 | | N(−4, 1) | N+(0, 1) |
| Cut-point 1 | | N(2, 1) | N+(0, 1) |
| **Covariates** | | | |
| Effect of age (threshold, RT intercept, confidence intercept) | | N(0, 0.3) | — |
| Effect of DPN (threshold, RT intercept, confidence intercept) | | N(0, 0.5) | — |

*Note.* N($\mu$, $\sigma$) denotes a normal distribution; N+ denotes a half-normal distribution (between-participant SDs are constrained to be positive). Participant-level deviations were standardised (z $\sim$ N(0, 1)), and correlations between participant-level parameters had an LKJ(2) prior. Thermal contrast entered the response time and confidence models on a 0–100 scale. Group-difference priors follow the pre-registration (N(0, 0.5)).
